# Hyperconnectivity and altered dynamic interactions of a nucleus accumbens network in post-stroke depression

**DOI:** 10.1101/2021.11.05.21265997

**Authors:** Lena KL Oestreich, Paul Wright, Michael J O’Sullivan

## Abstract

**Objectives:** Post-stroke depression (PSD) is a common complication after stroke. To date, no consistent locus of injury is associated with this complication. Here, we probed network dynamics in four functional circuits tightly linked to major depressive disorder and investigated structural alterations within these networks in PSD.

**Methods:** Forty-four participants with recent stroke and 16 healthy volunteers were imaged with 3T structural, diffusion and resting-state functional MRI and completed the Geriatric Depression Scale (GDS). Associations between GDS and functional connectivity were investigated within networks seeded from nucleus accumbens (NAc), amygdala and dorsolateral prefrontal cortex. In addition, the default mode network (DMN) was identified by connectivity with medial prefrontal cortex (mPFC) and posterior cingulate cortex (PCC). Circuits that exhibited altered activity associated with GDS were further investigated by extracting within-network volumetric and microstructural measures from structural images.

**Results:** Functional connectivity within the NAc-seeded network and DMN correlated positively with depressive symptoms. Normal anticorrelations between these two networks were absent in patients with PSD. PCC grey matter volume as well as microstructural measures in mPFC and the medial forebrain bundle, a major projection pathway interconnecting the NAc-seeded network and links to mPFC, were associated with GDS scores.

**Conclusions:** Depression after stroke is marked by reduced mutual inhibition between functional circuits involving NAc and DMN as well as volumetric and microstructural changes within these networks. Aberrant network dynamics present in patients with PSD are therefore likely to be influenced by secondary, pervasive alterations in grey and white matter, remote from the site of injury.

## INTRODUCTION

For approximately one-third of stroke survivors, rehabilitation outcomes are adversely affected by post-stroke depression (PSD).^1^ PSD is partly driven by mechanisms specific to brain injury rather than the psychological reaction to physical disability alone: stroke survivors exhibit higher rates of depression than physically ill patients with similar levels of disability.^2^ However, systematic reviews^3^ and meta-analyses^4^ have found no simple relationship between lesion size or location and PSD. One possible explanation is that infarction initiates a cascade of events, whereby alterations in brain structure and function propagate away from the site of injury, so that key alterations critical to behaviour come to be present outside the extent of visible injury on standard MRI. Advances in microstructural and functional MRI provide a methodology to characterise such alterations.

Studies in animal models of depression and major depressive disorder (MDD) in the absence of injury identify candidate networks that could also be implicated in the development of depression after stroke. The amygdala receives sensory input from thalamic projections^5^ and facilitates affective regulation in response to sensory stimuli via connections to dorsal anterior cingulate cortex (dACC) and insula.^6^ In MDD, hyperconnectivity accompanied by enlargement of amygdala has been observed.^7^ The nucleus accumbens (NAc) and associated networks are implicated by animal models of depression: the NAc receives dopaminergic input from the ventral tegmental area via the medial forebrain bundle, a major projection tract, that carries the mesolimbic dopamine pathway.^8^ Experimental inhibition of mesolimbic dopamine release leads to impaired defensive reactions during adverse conditions (learned helplessness) and blunted responses to reward (anhedonia),^9^ two cardinal features of depression in humans. The NAc modulates reward processing by integrating inputs from limbic and cortical regions including hippocampus, anterior cingulate and orbitofrontal cortex^10^ and also receives efferent projections from the amygdala.

Large-scale cortex-embedded networks have also been implicated in depressive symptoms in MDD, potentially via reduced top-down control of emotion processing.^11^ Limited responsiveness^12^ and smaller volumes^13^ of dorsolateral prefrontal cortex have been reported in MDD. Attenuated functional connectivity in this network has been observed 10 days^14^ and 3 months^15^ post-stroke. Another feature of studies in MDD is altered dynamic interaction between networks, for example between the salience network and DMN, leading to synchronous activation of both networks, which has been linked to difficulties for depressed patients to shift from self-focused thinking to goal-directed behavior.^16^ This type of reciprocal network interaction has not been investigated in stroke. One advantage of doing so is that the discovery of altered network dynamics, coinciding with emergence of depression, would provide stronger causal evidence for the role of network interactions in the development of depressive symptoms.

Based on the previous literature in animal models of depression and MDD, the primary aim of this study was to investigate whether functional networks typically affected in MDD without brain injury, also exhibit aberrant neural dynamics in individuals who experience depression after stroke. We further probed cross-network dynamics of these networks via correlation of activity with the DMN. A secondary aim was to explore mechanistic underpinnings of network dynamics associated with PSD. For this purpose, we conducted follow-up analyses in circuits exhibiting depression-related activity by testing properties of grey matter regions and white matter connections that provide their structural architecture.

## MATERIALS AND METHODS

### Participants

A sample of 46 patients with first ischemic stroke was recruited from a larger cohort enrolled into a longitudinal study of cognitive and behavioral features after stroke (STRATEGIC, NCT03982147). Participants were aged over 50 years and had a diagnosis of ischemic stroke confirmed by neuroimaging. Exclusion criteria were dementia, previous stroke, major neurological disease, previous moderate to severe head injury, inability to converse fluently in English, active malignancy, and any other factors that would affect performance of cognitive tasks (e.g., aphasia, visual impairment etc.). Participants also completed the Geriatric Depression Scale (GDS), a 30-item self-report measure designed to identify depression in older people.^17^ Sixteen age- and handedness-matched healthy controls were recruited from the community. The study protocol was approved by the London and Bromley Research Ethics Committee and the University of Queensland Research Ethics Committee. All participants gave written informed consent. Two participants were excluded due to motion artefacts in the resting-state functional MRI (rs-fMRI) scans.

### Data acquisition and processing

Research MRI was performed 30-95 days after stroke (*M* = 69.7, *SD* = 17.4) on a 3T MR750 MR scanner (GE Healthcare, Little Chalfont, Buckinghamshire, UK). T1-weighted images were acquired with the MPRAGE sequence^18^ using the following parameters: repetition time (TR) = 7.31ms, echo time (TE) = 3.02ms and flip angle = 11°. Images were acquired in the sagittal plane with field of view (FOV) = 270 × 270mm, matrix size = 256 × 256 voxels and slice thickness and gap = 1.2mm. T2-weighted fluid-attenuated inversion recovery (FLAIR) and fast recovery fast spin echo (FRFSE) sequences were acquired for lesion delineation. The FLAIR sequence was acquired with TR = 8000ms, TE = 120-130ms and flip angle = 90-111°. The FRFSE sequence was performed with TR = 4380ms, TE = 54-65ms and flip angle = 90-111°. Images were acquired in the axial plane with FOV = 240 × 240mm for both sequences. The matrix size for the FLAIR sequence was 256 × 128 voxels and 320 × 256 voxels for the FRFSE sequence. Slice positions were aligned for both sequences with 36 slices at 4mm thickness for FLAIR and 72 slices at 2mm thickness for FRFSE.

Rs-fMRI was acquired with an echo-planar imaging sequence using the following parameters: TR = 2000ms, TE = 30ms, flip angle = 75°, slice thickness = 3mm and gap = 0.3mm. Images were acquired in the axial plane with FOV = 211 × 211mm, matrix size = 64 × 64mm and number of slices = 40. A total of 180 volumes were collected over a scan time of 7min. Participants were instructed to keep their gaze on a fixation cross throughout the scan. Diffusion-weighted images were acquired with an echo-planar imaging sequence with double refocused spin echo for 60 diffusion-sensitization directions at *b*=1500s/mm^2^ and six acquisitions without diffusion sensitization (*b*=0). Image geometry for the diffusion-weighted images covered the whole brain using 2mm axial slices with matrix size of 128 × 128 voxels and FOV of 256 × 256mm, resulting in 2mm isotropic resolution. Slices were aligned such that the intercommissural line was as close to the axial plane as possible. Acquisition was peripherally gated to the cardiac cycle, giving a sequence duration of 11-20min, a TR of 10,000-14,118ms and a TE of 66-78ms with a flip angle of 90°.

### Rs-fMRI processing

Data were pre-processed and analyzed using the functional connectivity toolbox CONN version 19b (www.nitrc.org/projects/conn, RRID:SCR_009550),^19^ implemented in MATLAB R2020a and SPM12 (https://www.fil.ion.ucl.ac.uk/spm/software/spm12/). The first 10 volumes of each participant’s fMRI data were discarded to allow for magnetization equilibrium. After brain extraction, the T1-weighted images were centered (coordinates = 0, 0, 0), segmented and normalized. Images were smoothed with an 8mm fullwidth at half-maximum isotropic Gaussian kernel. Denoising was performed by utilizing the anatomical component-based noise correction procedure (CompCor), regressing noise components from cerebrospinal fluid and cerebral white matter,^20^ outlier scans or scrubbing identified during pre-processing,^21^ estimated subject-motion parameters,^22^ as well as constant and first-order linear session effects^19^ out of the fMRI time series. The data were then temporally bandpass-filtered (0.01 to 0.08Hz) to remove functional images with linear trends. Functional connectivity (FC) networks were defined based on seed regions in the nucleus accumbens (NAc-seeded network), amygdala (amygdala-seeded network) and dorsolateral prefrontal cortex (dlPFC-seeded network). The DMN was reconstructed by using seed regions in posterior cingulate cortex (PCC) and medial prefrontal cortex (mPFC). A visual network, seeded from the primary visual cortex (V1-seeded network), was used as a control network. Seeds were defined as spheres with a radius of 6mm (see Supplementary Figure 1), placed at the MNI coordinates of the NAc (left: −12, 08, −12; right: 10, 10, −12), amygdala (left: −24, 0, −21; right: 21, −1, −22), dlPFC (left: −46, 38, 8; right: 43, 38, 12), PCC (left: −39, 34, 37; right: 35, 39, 31), mPFC (left: −5, 17, −13; right: 7, 17, −14) and V1 (left: −11, −81, 7; right: 11, −78, 9). Individual FC maps were generated for each seed region, using averaged time series. The FC strength between seed region and all brain voxels was quantified by the Pearson’s cross-correlation. Fisher’s r-to-z transformation was applied to improve the normal distribution of the correlation coefficients. All analyses were FWE-corrected at a cluster threshold of p < 0.05.

### T1/DWI processing

T1-weighted images were preprocessed with the *recon-all* command implemented in FreeSurfer (v6.0) (http://surfer.nmr.mgh.harvard.edu/) to reconstruct cortical and subcortical parcellations based on the Desikan-Killiany atlas.^23^ Parcellations were validated by manual inspection. The diffusion-weighted data were corrected for head movements, eddy current distortions and magnetic field inhomogeneities, using tools implemented in MRtrix3.^24^ A five-tissue-type segmented image, suitable for use for anatomically constrained tractography, was generated from the pre-processed T1-weighted images. Free-water imaging was used to correct for CSF contamination and to quantify the amount of extracellular free-water (FW) by separating the diffusion properties of brain tissue from the surrounding extracellular free-water.^25^ Response functions were estimated using the single-fiber *tournier* algorithm^26^ and constrained spherical deconvolution was applied to obtain fiber orientation distributions (FOD). Anatomically constrained tractography with the 2^nd^ order integration over Fiber Orientation Distribution (iFOD2) algorithm,^24^ was used to generate individual tractograms for each participant.^27^ Tractograms were generated until 100 million streamlines were obtained with a length of 5-250mm, step size of 1mm and FOD amplitude threshold of 0.1. The spherical deconvolution informed filtering of tractograms (SIFT) algorithm was applied to reduce the overall streamline count to 10 million, providing more biologically meaningful estimates.^28^

### Follow-up analysis of volume and microstructure in functional networks associated with PSD

Volumes and extracellular free-water (FW) estimates were generated for relevant brain structures generated from FreeSurfer parcellations, namely NAc, mPFC and PCC. Clusters with significant FC to seed regions associated with GDS scores in any of the five networks were extracted as binary masks and registered to individual subject space. These masks, together with the original seed regions were then used as inclusion regions of interest (ROIs) through which tracts had to pass to be maintained from the original whole-brain tractograms. Free-water corrected (tissue-specific) fractional anisotropy (FA_t_) and FW were extracted and averaged across each tract. Fibers in the DMN could not be reconstructed for three participants from the D-group.

### Lesion definition and overlap with FC networks/ white matter tracts

FLAIR images were used to draw lesions manually. When necessary, acutely acquired diffusion images were used to identify the infarct. Lesion maps and white matter tracts were co-registered together with the rs-fMRI networks into MNI standard space, so that anatomically homologous brain areas were aligned. Overlap of resting-state networks, grey matter regions and white matter tracts with lesions were quantified with the Jaccard index, which was compared between groups and tested for associations with GDS scores.

### Statistical analysis

While most analyses were performed using GDS scores as a continuous variable, patients with stroke were split into groups of patients with PSD (D+) and patients free of PSD (D-) for some analyses to facilitate comparisons to the healthy control (HC) group. Stroke groups were split based on a score of 10 on the GDS, which has previously been identified to yield the highest sensitivity (0.69) and specificity (0.75) in stroke samples.^29^ Thirty-two (72.7%) stroke patients had a score below 10 on the GDS and were assigned to the D-group and 12 (27.3%) stroke patients had a score of 10 or above on the GDS and were assigned to the D+ group.

To investigate whether FC in any of the five networks was associated with PSD, GDS scores of participants in the stroke group were regressed onto the whole-brain FC maps estimated from each of the seed regions (NAc/ amygdala/ dlPFC/ PCC/ mPFC/ V1). Average FC was then extracted from clusters that were significantly associated with GDS scores, individually for each participant. A repeated-measures analysis of covariances (ANCOVA) with group (HC/D+/D-) as between-subjects factor and network and cluster as within-subjects factors was calculated to determine if FC in these clusters varied across groups. Several regions of the identified resting-state networks are anticorrelated with one another (see Supplementary Figure 1). To investigate whether FC changes associated with PSD are driven by insufficient deactivation of anticorrelated networks, a multivariate analysis of covariances (MANCOVA) with group (HC/D+/D-) as between-subjects factor and average FC (extracted from anticorrelated clusters) as within-subjects factor was performed.

### Follow-up statistical analysis of volume and microstructure in functional networks associated with PSD

Two repeated-measures ANCOVA with group (HC/D+/D-) as between-subjects factor, as well as volume or FW (NAc/PCC/mPFC) and hemisphere (left/right) as within-subjects factors were conducted to test for structural alterations in the seed regions between groups. To determine whether BOLD time-series correlations between seed regions and clusters associated with PSD were accompanied by underlying changes in white matter microstructure, Pearson’s correlation coefficients between GDS scores and microstructural measures (FA_t_/FW) were calculated in the stroke sample only. Group differences were tested using a MANCOVA with the between-subjects factor group (HC/D-/D+) and the within-subjects factors white matter estimates (FA_t_/FW).

Age and sex were added as covariates to all analyses. All p-values were adjusted using the Bonferroni correction of *p = α/k*, whereby alpha was set to 0.05 and *k* denoted the number of comparisons.

## RESULTS

Stroke and HC groups did not differ in age or handedness, but there were more men in the stroke group than in the HC group (see Table 1). The D+ and D-groups had a similar gender ratio of approximately 1:3 women to men (*X*^*2*^ (1) = 0.05, *p* = 0.826).

**Table 1.** Demographics and lesion characteristics by group.

| Variable | HC (n = 16)<br>mean (SD)/ category | D+ (n = 12)<br>mean (SD)/ category | D- (n = 32)<br>mean (SD)/ category | group comparisons |
| --- | --- | --- | --- | --- |
| Demographics |  |  |  |  |
| Age (years) | 71.53 (10.62) | 69.72 (6.96) | 68.78 (9.22) | $F(2,57) = 0.48, p = 0.62$ |
| Sex (female/male) | 11/5 | 3/9 | 7/25 | $\chi^2(2) = 10.96, p = 0.004$ |
| Handedness (right/left) | 16/0 | 11/1 | 31/1 | $\chi^2(2) = 1.49, p = 0.48$ |
| Lesion characteristics |  |  |  |  |
| Hemisphere (left/right/bilateral) | | 5/7/0 | 18/13/1 | $\chi^2(2) = 1.33, p = 0.51$ |
| Arterial territory (ACA/MCA<sub>ant</sub>/MCA<sub>pos</sub>/MCA<sub>str</sub>/PCA/lacunar/thalamic) | | 0/2/3/3/2/1/1 | 1/7/7/4/6/5/2 | $\chi^2(6) = 1.80, p = 0.94$ |
| Volume (ml) | | 7175.64 (12081.03) | 7600.81 (11962.09) | $t(42) = 0.01, p = 0.92$ |
| Time since lesion (days) | | 68.17 (13.39) | 70.28 (18.85) | $t(42) = 0.13, p = 0.72$ |

Note. HC = healthy controls; D+ = Post-stroke depression group; D- = stroke group free of depression; SD = standard deviation; MCA<sub>ant</sub> = middle cerebral artery, anterior; ACA = anterior cerebral artery; MCA<sub>pos</sub> = middle cerebral artery, posterior; MCA<sub>str</sub> = middle cerebral artery, striatocapsular; PCA = posterior cerebral artery

### Seed-based resting-state networks

The five seeded FC networks averaged across all participants and their locations are shown in Supplementary Figure 1 and their cluster size and locations are provided in Supplementary Table 1. Briefly, a positive BOLD time-series correlation was found between NAc and a single large cluster largely confined to adjacent subcortical structures but extending to insula and medial frontal cortex (including parts of the dorsal and ventral ACC as well as orbitofrontal cortex). The amygdala-seeded network subsumed similar subcortical structures and medial frontal regions but also involved parts of the temporal lobe (parahippocampal gyrus, middle temporal gyrus, superior temporal gyrus, temporal pole) and lingual gyrus. The dlPFC-seeded network included bilateral clusters including the frontal pole, inferior frontal gyrus and superior frontal gyrus, bilateral clusters spanning across regions in the lateral occipital cortex, angular gyrus and supramarginal gyrus and a medial cluster including dorsal ACC and superior frontal gyri. The DMN and V1-seeded networks corresponded with the typical configuration of these networks described in previous literature (Supplementary Figure 1). Supplementary Figure 1 also shows regions anticorrelated with each of these networks. Of note, BOLD activity in the NAc network was anticorrelated with a region of posterior cingulate cortex that was part of a larger region within the DMN.

### Associations between PSD and resting-state networks

Within the stroke group, GDS scores were associated with BOLD signal correlations between NAc and one cluster in the left (*t*(43) = 4.65, *p* = 0.001, *k* = 792) and one in the right (*t*(43) = 4.64, *p* = 0.004, *k* = 607) hemisphere (see Figure 1). Both clusters included the putamen, amygdala, orbitofrontal cortex and insula. The cluster seeded from the right NAc additionally covered areas of the pallidum and hippocampus in the right hemisphere. Within the DMN, GDS scores were associated with BOLD time-course correlations between PCC and a cluster including the medial frontal cortex and frontal poles (*t*(43) = 6.76, *p* < 0.001, *k* = 3789). GDS scores were furthermore associated with BOLD time-course correlations between the mPFC and a cluster in the precuneus and PCC (*t*(43) = 5.56, *p* < 0.001, *k* = 3251). In all four identified clusters, GDS increased as FC increased, indicating that hyperconnectivity within the NAc-seeded network and the DMN are associated with depression severity in stroke patients. No associations between GDS scores and FC in the amygdala-, dlPFC-, or V1-seeded networks were observed.

**Figure 1.**
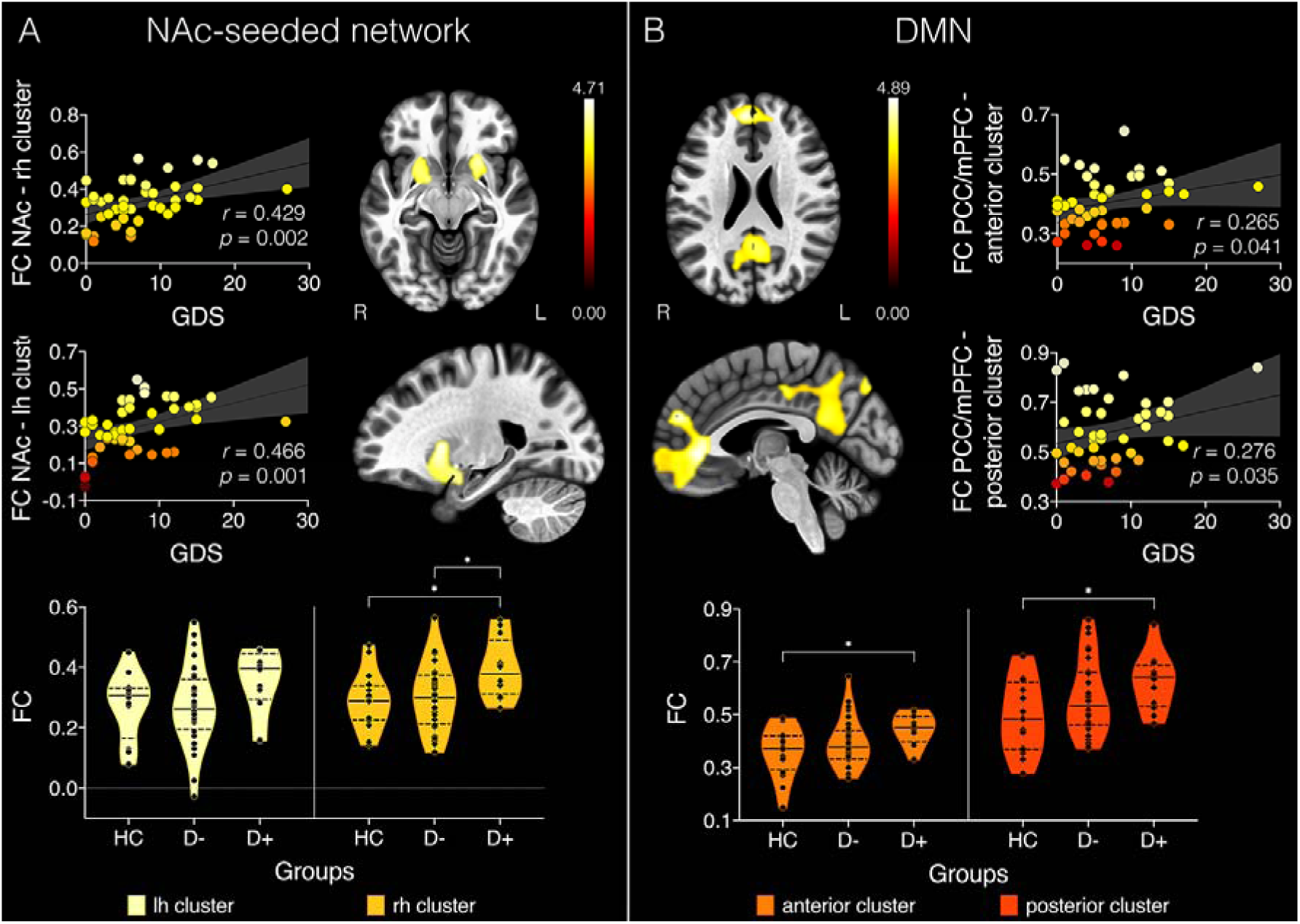
Clusters with significant BOLD time-series correlations with geriatric depression scale (GDS) (A) in nucleus accumbens (NAc)-seeded network and (B) the default mode network (DMN). Colorbars represent t-statistic values and data points in scatterplots are color-coded according to their corresponding t-statistic. FC = functional connectivity; PCC = posterior cingulate cortex; mPFC = medial prefrontal cortex; rh = right hemisphere; lh = left hemisphere; R = right; L = left; HC = healthy controls; D- = stroke patients free of depression; D+ = patients with post-stroke depression. Violin plots represent the distributions of FC values per group by cluster. \**p* < 0.05.

Average FC was extracted from each of the four clusters associated with GDS scores individually for each participant. A repeated-measures ANCOVA identified a significant main effect of group (*F*(2,55) = 6.25, *p* = 0.004, *n*_*p*_^*2*^ = 0.183). As can be seen in Figure 1, Bonferroni corrected post-hoc tests (12 comparisons) found that FC between the right hemisphere cluster and NAc was significantly increased in the D+ group compared to the HC group (*t*(26) = 2.70, *p* = 0.027) and the D-group (*t*(42) = 2.68, *p* = 0.029). Furthermore, FC in clusters correlated with the DMN seeds was significantly increased in the D+ group compared to the HC group (anterior cluster: *t*(26) = 2.52, *p* = 0.044; posterior cluster: *t*(26) = 2.51, *p* = 0.045).

### Anticorrelations between DMN and NAc-seeded network

Only the NAc-seeded network exhibited significant anticorrelations with regions in the DMN. For every participant, average FC was extracted from clusters that were anticorrelated between DMN and NAc-seeded network. A MANCOVA identified main effects of group for FC between bilateral NAc and PCC/precuneus (*F*(4,55) = 11.8, *p* < 0.001, *n*_*p*_^*2*^ = 0.293), as well as PCC and thalamus (*F*(4,55) = 10.44, *p* < 0.001, *n*_*p*_^*2*^ = 0.268) and mPFC and thalamus (*F*(4,55) = 4.75, *p* = 0.012, *n*_*p*_^*2*^ = 0.143) (see Figure 2). Specifically, Bonferroni corrected post-hoc tests (12 comparisons) identified significantly decreased FC (i.e. a greater degree of anticorrelation) in the HC group compared to the D+ group in all three clusters (bilateral NAc – PCC/precuneus: *t*(26) = 4.68, *p* < 0.001; PCC – thalamus: *t*(26) = 4.57, *p* < 0.001; mPFC – thalamus: *t*(26) = 2.86, *p* = 0.018). The D+ group also exhibited reduced anticorrelated FC compared to the D-group between the bilateral NAc and PCC/precuneus (*t*(42) = 4.01, *p* = 0.001) and PCC and thalamus (*t*(42) = 2.81, *p* = 0.02) clusters. Lastly, the D-group had significantly increased FC compared to the HC between PCC and thalamus (*t*(46) = 2.59, *p* = 0.037) and mPFC and thalamus (*t*(46) = 2.5, *p* = 0.046). As can be seen in Figure 2, on average, FC values changed from negative to positive in the D+ group for all three clusters, indicating that DMN activity in these areas is not adequately suppressed during activation of the NAc-seeded network and vice versa.

**Figure 2.**
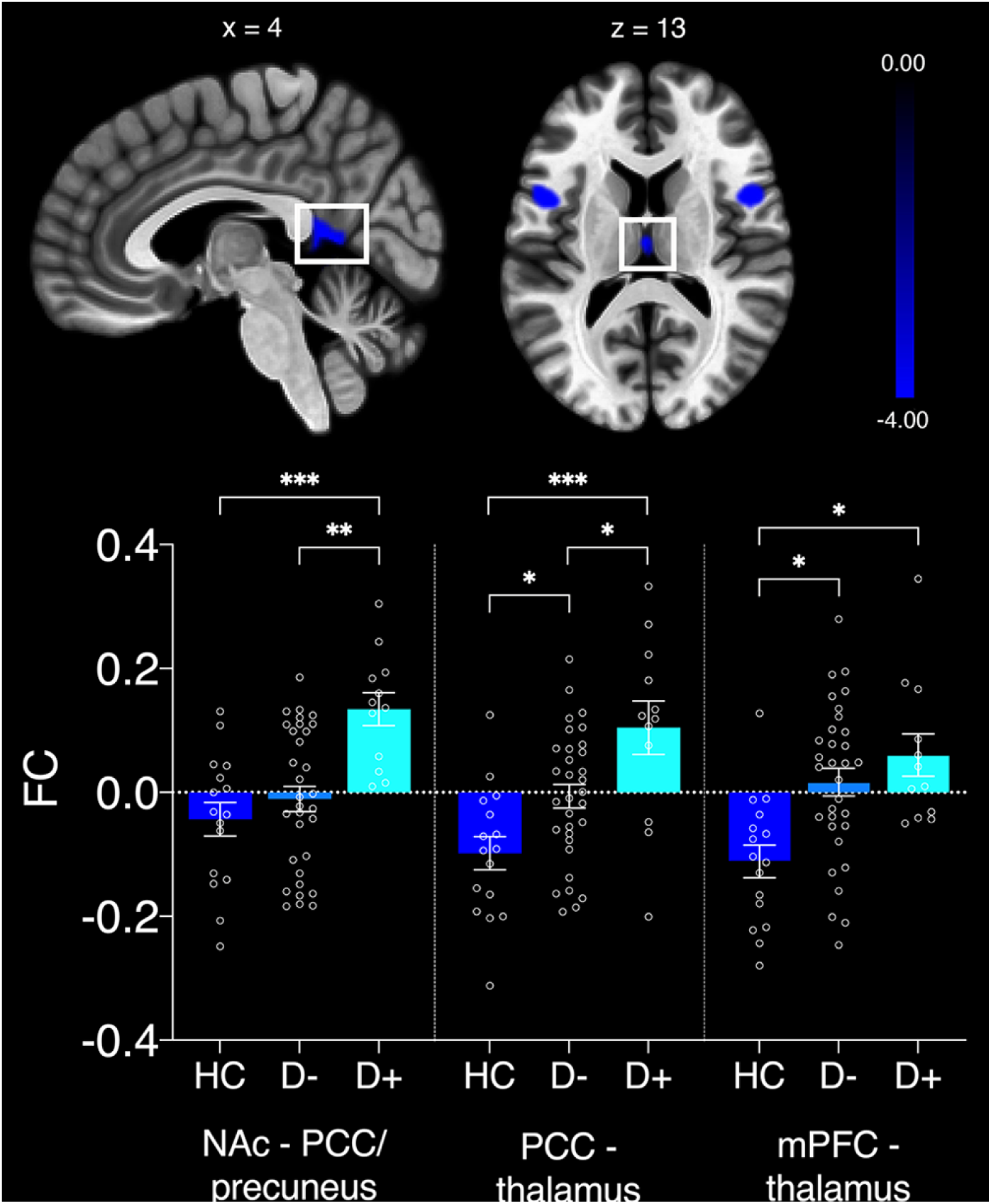
Significant group differences in anticorrelated clusters of nucleus accumbens (NAc)-seeded network and default mode network (DMN). FC = functional connectivity; PCC = posterior cingulate cortex; mPFC = medial prefrontal cortex. Error bars represent standard errors of the mean. \**p* < 0.05; \*\**p* < 0.01; \*\*\**p* < 0.001.

### Follow-up analysis of volume and microstructure in functional networks associated with PSD: NAc-seeded network and DMN

#### Grey matter volume and microstructure

After Bonferroni correction for 12 comparisons, volume of the right PCC was negatively correlated with GDS scores (*r* = −0.286, *p* = 0.03), whereby volume decreased as depression severity increased (see Figure 3). FW correlated positively with GDS scores in the right PCC (*r* = 0.4, *p* = 0.024), left (*r* = 0.3, *p* = 0.048) and right mPFC (*r* = 0.312, *p* = 0.039). This indicates that FW increases with depression severity in the seed regions of the DMN. While no significant main effects or interactions were found with volume, a repeated-measures ANCOVA with FW identified a main effect of group (*F*(2,55) = 5.52, *p* = 0.006, *n*_*p*_^*2*^ = 0.162). Bonferroni corrected post-hoc tests (9 comparisons) identified increased FW in the D+ group compared to the HC group in the left NAc (*t*(26) = 3.13, *p* = 0.036), right PCC (*t*(26) = 3.24, *p* = 0.039), left mPFC (*t*(26) = 2.79, *p* = 0.019) and right mPFC (*t*(26) = 3.46, *p* = 0.027). Increased FW was also observed in the D+ group compared to the D-group in the right PCC (*t*(42) = 2.98, *p* = 0.014) and left mPFC (*t*(42) = 3.43, *p* = 0.027).

**Figure 3.**
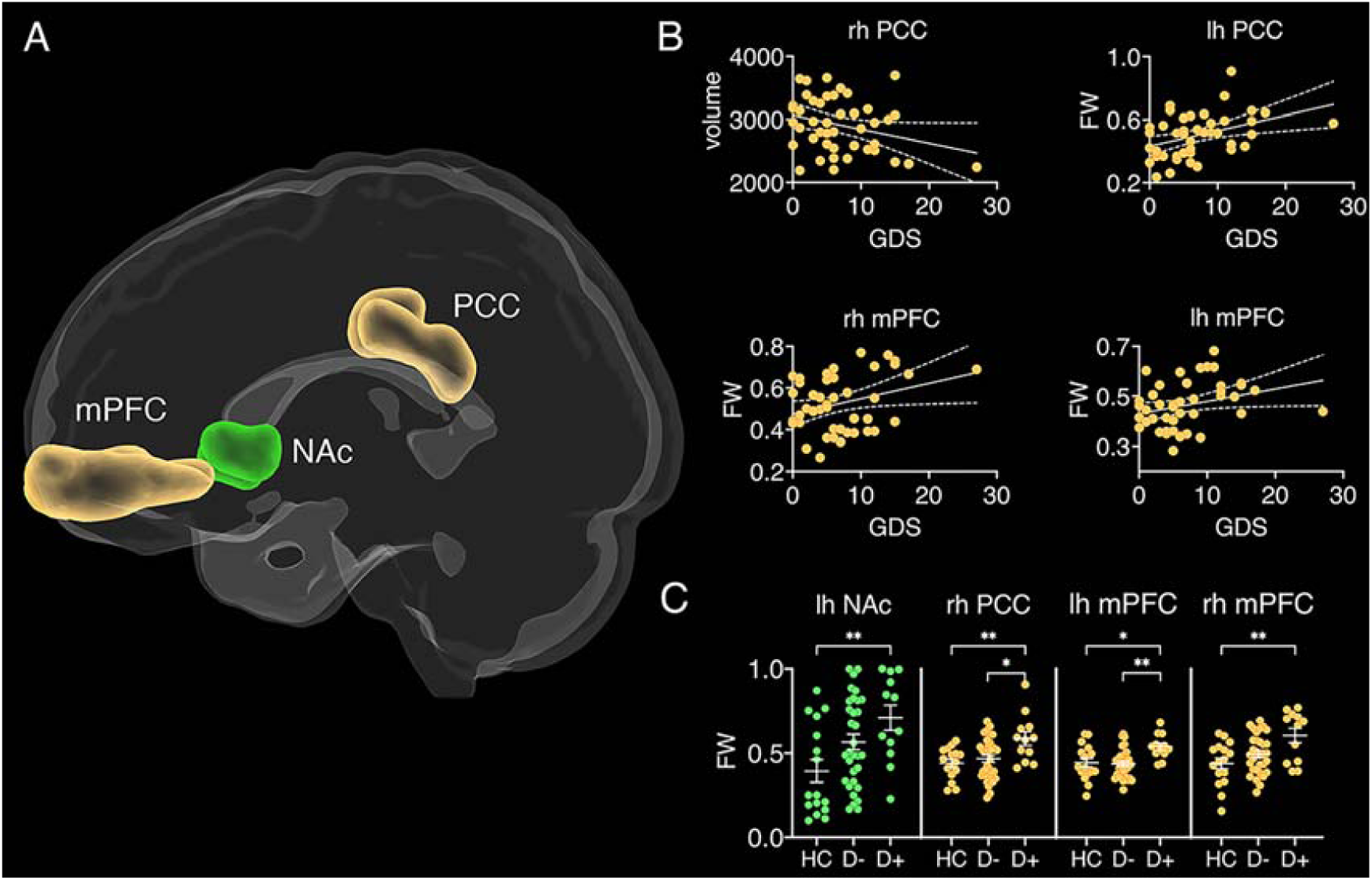
Grey matter seed regions of resting-state networks. (A) dlPFC = dorsolateral prefrontal cortex (teal), PCC = posterior cingulate cortex (yellow), mPFC = medial prefrontal cortex (yellow), NAc = nucleus accumbens (green), Amy = amygdala (light blue), V1 = primary visual cortex (blue). (B) correlations between geriatric depression scale (GDS) scores and free-water (FW)/volume of grey matter regions. Error bands represent standard error, rh = right hemisphere, lh = left hemisphere. (C) Group differences in FW for different grey matter structures. HC = healthy controls, D+ = stroke patients with depression, D- = stroke patients free of depression.; \**p* < 0.05; \*\**p* < 0.01; \*\*\**p* < 0.001.

#### White matter tract microstructure

Tractography analysis suggested that the majority of fibers connecting NAc to the positively functionally correlated subcortical and frontal lobe cluster coursed in the medial forebrain bundle. Connection streamlines between ROIs in the DMN were located in the dorsal cingulum bundle (see Figure 4). After Bonferroni-correction for 4 comparisons, FA_t_ correlated negatively with GDS scores in both tracts (medial forebrain bundle: *r* = −0.279, *p* = 0.033; dorsal cingulum bundle: *r* = −0.319, *p* = 0.021). In both cases, FA_t_ decreased with increasing depression severity. FW correlated positively (*r* = 0.450, *p* = 0.003) with GDS scores in tracts interconnecting the NAc-seeded network (i.e., medial forebrain bundle), whereby FW increased with increasing depression severity. A MANCOVA identified significant main effects for FA_t_ (medial forebrain bundle: *F*(4,55) = 7.91, *p* = 0.001, *n*_*p*_^*2*^ = 0.23; dorsal cingulum bundle: *F*(4,55) = 3.63, *p* = 0.033, *n*_*p*_^*2*^ = 0.119) and FW (medial forebrain bundle: *F*(4,55) = 4.26, *p* = 0.019, *n*_*p*_^*2*^ = 0.139; dorsal cingulum bundle: *F*(4,55) = 4.29, *p* = 0.019, *n*_*p*_^*2*^ = 0.139). Bonferroni corrected post-hoc tests (12 comparisons) revealed that FA_t_ was significantly reduced and FW significantly increased in the D+ group compared to the HC group in fibers of the NAc-seeded network, i.e., the medial forebrain bundle (FA_t_ : *t*(26) = 3.97, *p* = 0.012; FW: *t*(26) = 2.84, *p* = 0.019) and tracts interconnecting the DMN, i.e., the dorsal cingulum bundle (FA_t_ : *t*(26) = 2.67, *p* = 0.03; FW: *t*(26) = 2.89, p = 0.017).

**Figure 4.**
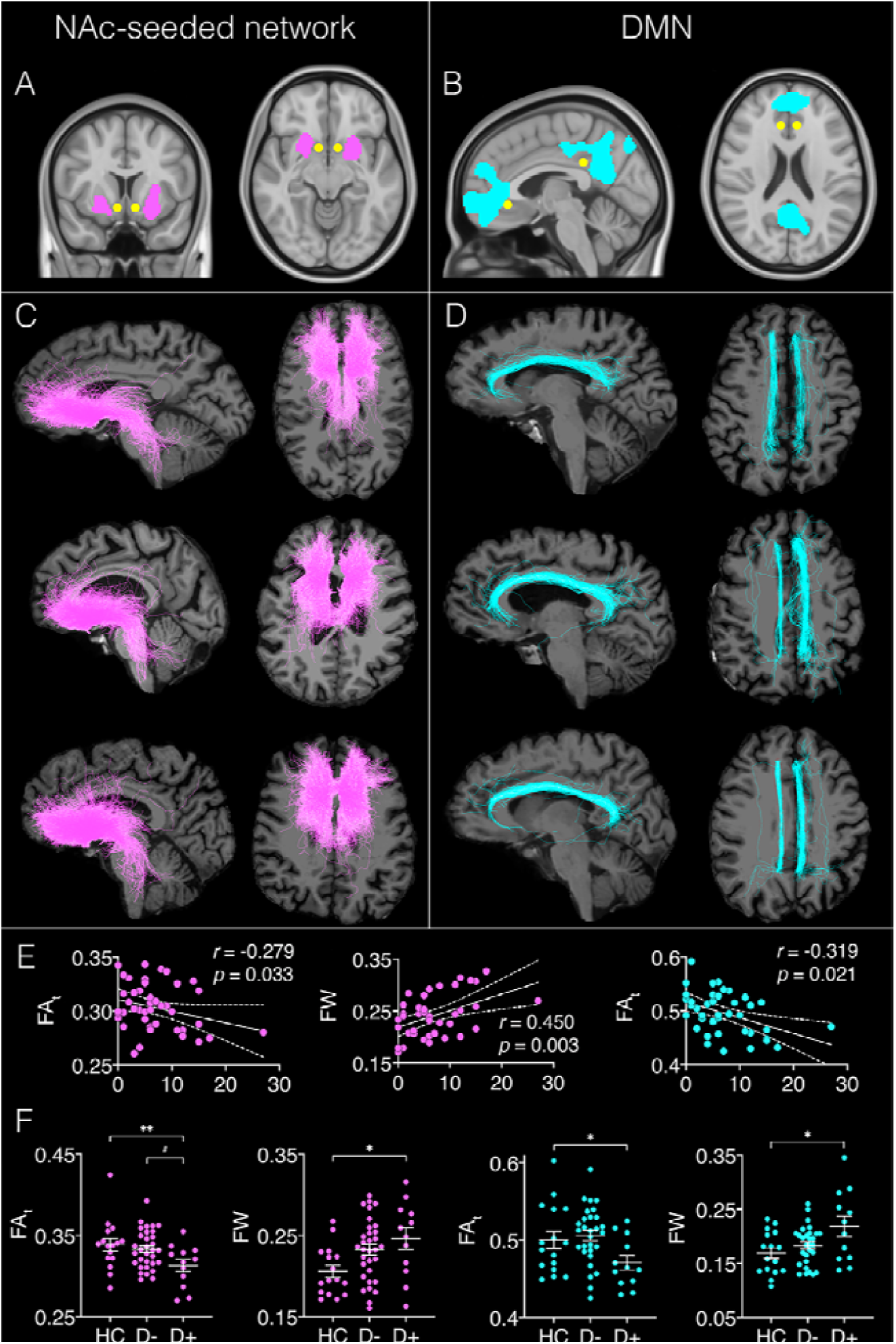
Regions of interest (ROIs) used for tractography and reconstructed white matter tracts. Inclusion ROIs are (A) nucleus accumbens (NAc) seed (yellow) and cluster with significant BOLD time series correlations associated with geriatric depression scale (GDS) scores (pink); and (B) posterior cingulate cortex (PCC) and medial prefrontal cortex (mPFC) seeds (yellow) of the default mode network (DMN) and cluster with significant BOLD time series correlations associated with GDS scores (blue); (C) Example of white matter tracts reconstructed for three participants using the ROIs of the NAc-seeded network, warped to individual subject space. The majority of fibers map onto the medial forebrain bundle. (D) Example of white matter tracts from three participants using the ROIs of the DMN, warped to individual subject space. The majority of fibers map onto the dorsal cingulum bundle. (E) significant correlations between FA_t_/FW and GDS scores in the medial forebrain bundle (pink) and dorsal cingulum bundle (blue). F) Significant group differences in FA_t_/FW for the medial forebrain bundle (pink) and dorsal cingulum bundle (blue). HC = healthy controls, D+ = stroke patients with depression, D- = stroke patients free of depression. #*p* < 0.075; \**p* < 0.05; \*\**p* < 0.01; \*\*\**p* < 0.001.

#### Lesion overlap with FC networks, clusters associated with GDS scores, and white matter tracts

Lesion overlap with networks, tracts and clusters ranged from 0.02% to 1.28%. No significant group differences or associations with GDS scores were observed for Jaccard indices or voxel overlap with lesions (see Supplementary Figure 2 and Supplementary Table 2).

## DISCUSSION

Investigation of functional networks in a post-stroke cohort led to three main findings. Depressive symptoms were associated with hyperconnectivity in two functional networks: one seeded from NAc; and the DMN. Secondly, anticorrelation of activity between NAc and DMN diminished with increasing depressive symptoms, indicating altered interactions between these networks. In patients reaching criteria for PSD, anticorrelation was lost entirely. Mutual inhibition between task-associated networks and DMN is a characteristic feature of large-scale neural dynamics in the human brain. The current data show that in PSD, this mutual inhibition is lost between a network involved in reward processing and the DMN. Finally, volumetric and microstructural alterations were found within relevant networks suggesting that alterations in functional dynamics reflect pervasive structural change after stroke.

Disturbance of reward processing, through altered function in a network of regions linked to NAc, is a plausible mechanism for the emergence of low mood after stroke. BOLD activity in NAc was positively correlated with activity in a network of regions which subsumed the basal ganglia and limbic system and extended to medial frontal cortex, ACC, and orbitofrontal cortex. To our knowledge, this is the first study to identify enhanced functional activity in a network implicated in reward^30^ as associated with PSD. Functional aberrations in this network have been reported in MDD and late-life depression, where they have been linked to enhanced reactivity to punishment and increased suicidality.^31^ Our findings therefore suggest that depressive symptoms may share a mechanism of altered reward processing that is common between PSD and other depressive disorders. Previous studies of anticorrelation between DMN and the salience network have posited an inability to shift from self-focused thinking to goal-directed behavior as a behavioral consequence of reduced network interactions.^16^ An inability to shift to reward-directed behavior could also contribute to depressive symptoms, particularly behavioral apathy.

Hyperconnectivity in the DMN is a common observation across depressive disorders, including PSD.^32^ It has been suggested that rumination – a key feature of depression – is associated with excessive DMN activity, which in turn is thought to exacerbate switching out of self-reflective states in response to external demands.^33^ Interestingly, our findings reveal a connection between hyperconnectivity in the DMN and activity in the NAc-seeded network: PSD patients exhibited a reversal of anticorrelation between these two networks. During externally, goal-directed attention, DMN activity reduces in a load-depended fashion as cognitive demands increase,^34^ such that activity in the DMN decreases with increasing activity in NAc-seeded reward circuits. Greater anticorrelation between the two systems is linked to more efficient value-guided behavior and reward attitudes in humans,^35^ which indicates that coupling strength of the two networks is an important factor in successful application of reward processing in goal-directed behavior. NAc plays a central role in the pathophysiology of reward deficits and inefficient interactions with the DMN are thought to be implicated in the manifestation of anhedonia^36^ across mood and psychotic disorders. A study by Sharma et al. (2017) investigated reward responsivity in patients with MDD,^37^ bipolar disorder, schizophrenia, and healthy controls. The study observed decoupling between NAc and DMN regions with increasing reward sensitivity, and hyperconnectivity within the DMN, to be associated with reward deficits across all clinical groups relative to healthy controls. This lack of integration between DMN and NAc activity was interpreted to reflect a brain phenotype of impaired reward-oriented internal cognition, expressed externally as anhedonia. A failure of efficient communication between these two networks may therefore represent a cross-diagnostic neurological signature of reward-related depressive symptoms.

Impaired interaction between reward-processing and DMN in the brain provides a plausible causal mechanism for the emergence of depressive symptoms after stroke. Such an account would also provide an explanation for the absence of a simple consistent locus for depression in lesion-mapping studies. Correlation of BOLD signal variation suggests that NAc is functionally interconnected with multiple subcortical structures as well as regions of medial frontal and temporal cortex. Similarly, the DMN spans a distributed network of regions in frontal and parietal cortices. Consequently, structural lesions in a wide range of locations could potentially disturb both within-network and between-network functional interactions.

Microstructure of the medial forebrain bundle was associated with GDS scores. The medial forebrain bundle is implicated in reward processing through its connectivity with structures known to be critical for reward, and through carriage of the mesolimbic dopaminergic pathway from the ventral tegmental area to NAc.^38^ Furthermore, axonal tracing studies in rodents and non-human primates have shown that it carries direct connections to mPFC.^39, 40^ It is therefore a candidate pathway for functional interactions between NAc, regions of the mPFC involved in reward processing, and potentially other networks that involve mPFC including the DMN. Consistent with this account, microstructure of mPFC regions correlated with depressive symptoms, in a way that paralleled the associations found for microstructure of the medial forebrain bundle.

Structural changes were also found within the DMN, in conjunction with hyperconnectivity demonstrated in this network. Lower grey matter volume in the PCC was associated with higher depression scores. Volume reduction in PCC has previously been reported in patients with MDD and has been linked to altered DMN activity.^13^ The PCC is one of the most metabolically active brain regions and has dense structural connections to widespread regions of the brain.^41^ As part of the DMN, the PCC is most active during self-referential processes, but it is simultaneously the main cortical hub responsible for switching between DMN and other functional network activity in response to environmental changes requiring behavioral adaptation.^41^ It is possible that morphometric changes of PCC play a role in the loss of efficient cross-network communication between DMN and the NAc-seeded reward network, in patients with PSD. Microstructure of the dorsal cingulum bundle, which contains projections connecting PCC and precuneus to medial prefrontal cortex in the DMN,^42, 43^ also exhibited microstructural properties associated with depression severity in stroke survivors.

The basis for the structural alterations associated with depressive symptoms in some individuals remains unclear. The brain undergoes dynamic changes in the weeks to months after stroke, which extend beyond the initial site of injury. Microglial activation, for example, is known to propagate along white matter pathways away from a site of infarction,^44, 45^ and elevation of extracellular free water – as demonstrated in the medial forebrain bundle and dorsal cingulum in this study – has been considered a putative marker of neuroinflammation. This account, however, remains speculative. The main alternative explanation is that a structural pattern of vulnerability to depressive symptoms is present prior to stroke and that the interaction between stroke and pre-existing vulnerability precipitates the emergence of depressive symptoms.

No association was found between functional connectivity in amygdala and dlPFC-seeded networks and depressive symptoms in PSD. Functional changes in amygdala-seeded structures have previously been linked to increased cortisol levels in patients with MDD and patients with bipolar disorder who experienced depressive episodes.^7^ Cortisol levels are elevated in the first few days to weeks after stroke,^46^ which may explain why studies by Zhang et al. (2014, 2018) observed hyperconnectivity in this network in patients with PSD within the first 10 days of stroke. The current study was conducted approximately three months after stroke, when cortisol levels are typically restored to the normal reference range.^46^ Aberrant signaling in cortico-cortical circuits are commonly observed in healthy individuals with a family history of depression^47^ and in patients with MDD who achieve remission from depressive symptoms, but continue to experience cognitive impairments.^33^ It is possible that depression experienced during different stages of stroke recovery may be caused by different underlying neural mechanisms.

Cerebrovascular pathology is often found to coexist with late-life depression, so that the boundaries of these two disorders converge in older people. There is a need for future studies to investigate etiological brain-based mechanisms of depression across varying ages and in the context of neurological and vascular disorders. A methodological limitation of the current study was that diffusion MRI acquisition was limited to a single shell, so that more advanced compartment models could not be applied.

Collectively, the findings from this study suggest that aberrant activity within a network involving the NAc, specialized for processing signals linked to reward, and interaction between this network and the DMN, may give rise to depressive symptoms after stroke. Longitudinal studies are now needed to track the emergence and persistence of these structural and functional features after stroke. Use of other imaging modalities, blood and CSF biomarkers will help to elucidate the nature of these changes and investigate a putative role for neuroinflammation, similar to that proposed for MDD in the absence of stroke.

## Data Availability

All data produced in the present study are available upon reasonable request to the authors

## ACKNOWLEDGEMENTS

The authors thank the coordinators of the King’s Hyperacute Stroke Research Centre for their help with identifying and recruiting patients, and the manager and staff of the NIHR Wellcome Trust King’s Clinical Research Facility. The authors thank Ofer Pasternak for the provision of the code required for the free-water analysis.

## DATA SHARING

The data presented in this study will be made available upon request. Requests should be directed to the chief investigator (MJO), as defined in the study protocol.

