## Supplementary figures and images for "Hyperconnectivity and altered dynamic interactions of a nucleus accumbens network in post-stroke depression"

### Supplementary Figure 1

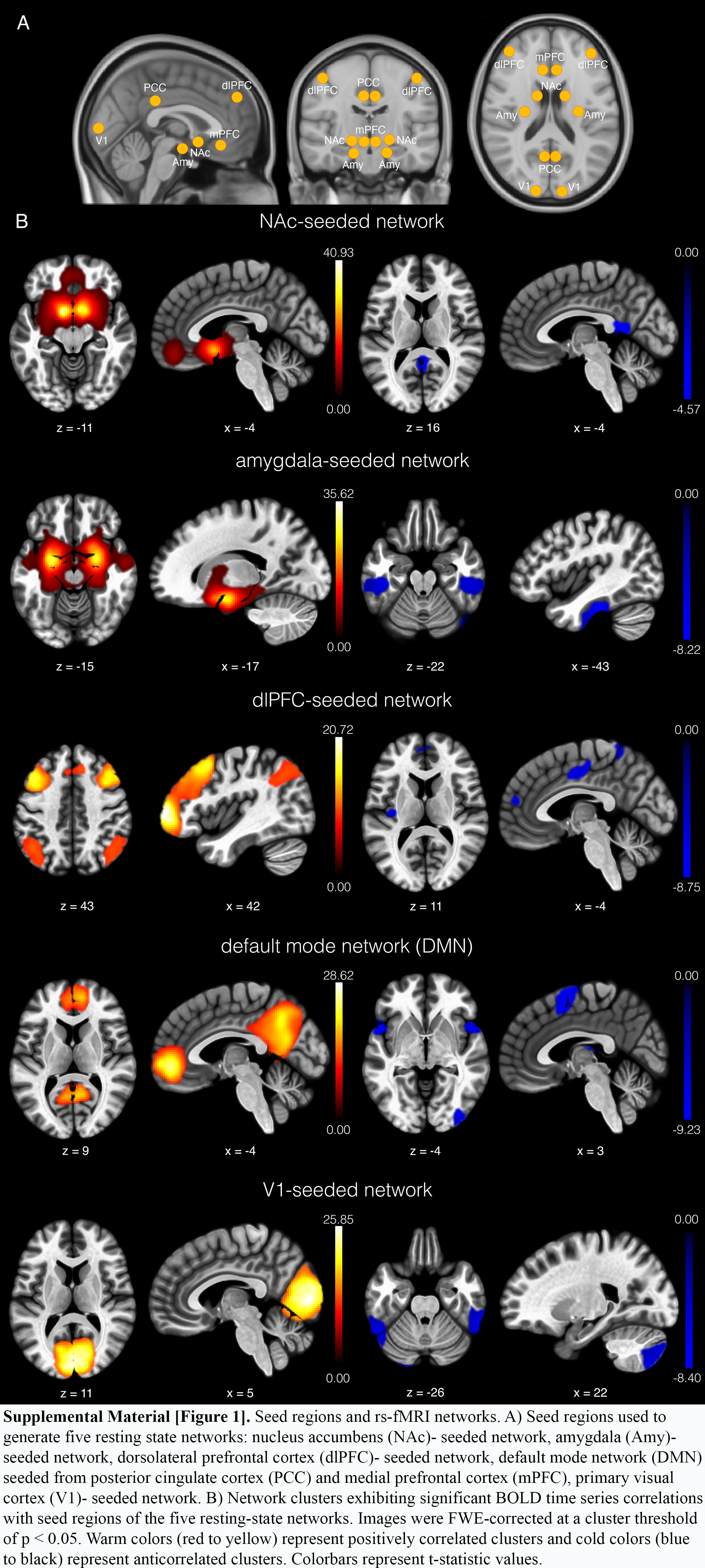

### Supplementary Figure 2

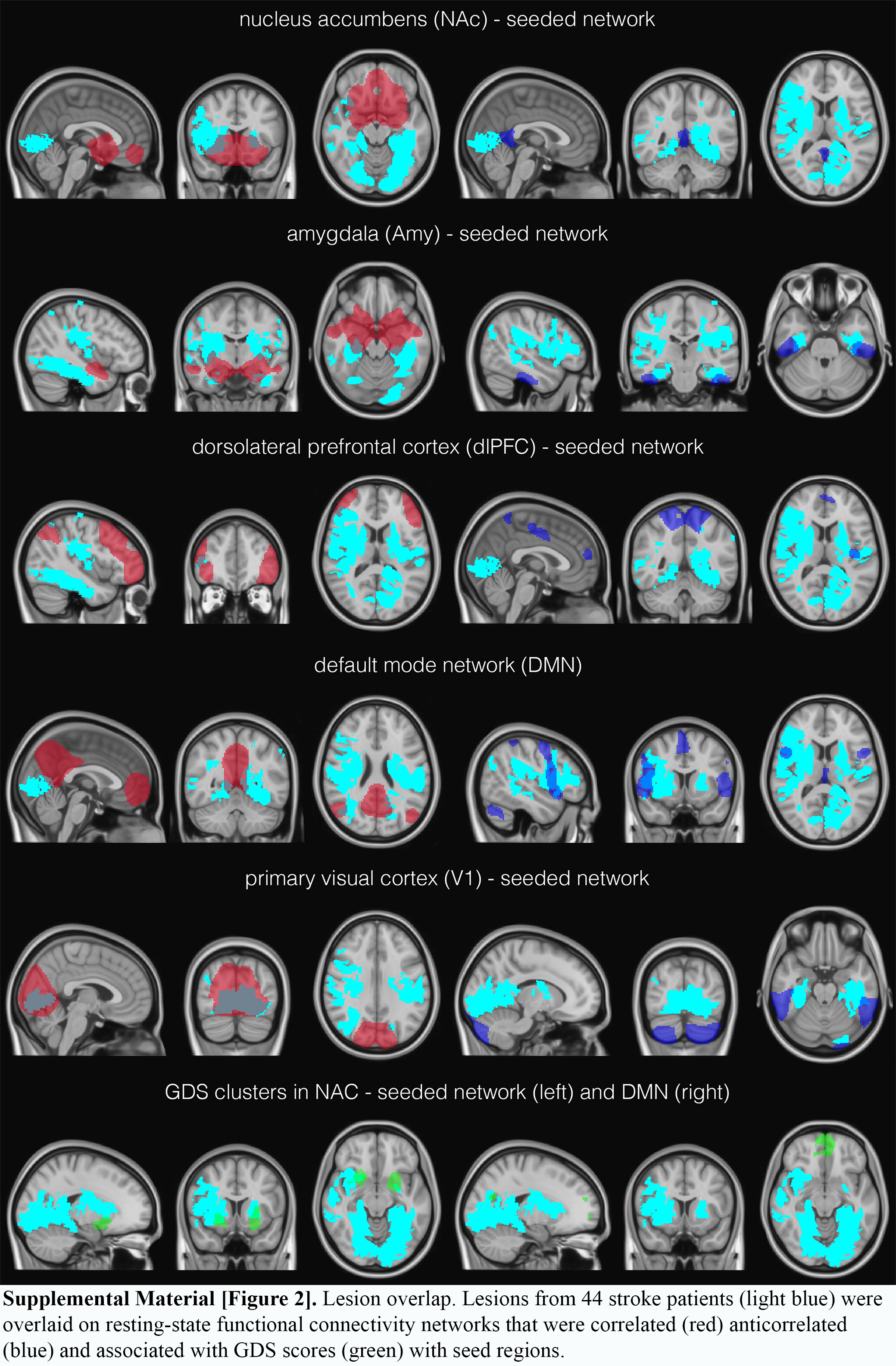
