## Supplementary Table 1 for "Hyperconnectivity and altered dynamic interactions of a nucleus accumbens network in post-stroke depression"

| Supplementary Table 1. Clusters correlated/anticorrelated with seed-regions | | | |
|---|---|---|---|
| | max intensity voxel | cluster size (voxels) | cluster location (number of voxels) |
| *NAc-seeded network – correlations* | | | |
| cluster 1 | –26 +28 +00 | 13 767 | left orbitofrontal cortex (1000) |
| | | | ventral anterior cingulate cortex (910) |
| | | | right orbitofrontal cortex (881) |
| | | | medial frontal cortex (689) |
| | | | left putamen (575) |
| | | | right putamen (476) |
| | | | left insular (370) |
| | | | left amygdala (321) |
| | | | right amygdala (291) |
| | | | left dorsal anterior cingulate cortex (280) |
| | | | right insular cortex (260) |
| | | | left caudate (258) |
| | | | right caudate (230) |
| | | | left pallidum (161) |
| | | | right dorsal anterior cingulate cortex (147) |
| | | | left thalamus (146) |
| | | | right thalamus (126) |
| | | | right pallidum (124) |
| | | | right hippocampus (106) |
| | | | left hippocampus (75) |
| | | | anterior cingulate gyrus (64) |
| *NAc-seeded network – anticorrelations* | | | |
| cluster 1 | +02 –50 +06 | 820 | posterior cingulate cortex (358) |
| | | | precuneous (244) |
| *Amy-seeded network – correlations* | | | |
| cluster 1 | –26 –06 –20 | 12 519 | right putamen (684) |
| | | | left putamen (656) |
| | | | left hippocampus (639) |
| | | | right parahippocampal gyrus (603) |
| | | | middle temporal gyrus (600) |
| | | | right hippocampus (562) |
| | | | left parahippocampal gyrus (554) |
| | | | right orbitofrontal cortex (447) |
| | | | left orbitofrontal cortex (421) |
| | | | left temporal pole (314) |
| | | | right temporal pole (300) |
| | | | right superior temporal gyrus (263) |
| | | | right insular (226) |
| | | | left insular (222) |
| | | | ventral anterior cingulate cortex (109) |

| | | | |
|---|---|---|---|
| | | | left pallidum (106)<br>right pallidum (103)<br>left planum polare (69)<br>right planum polare (55)<br>left superior temporal gyrus (51)<br>left nucleus accumbens (40)<br>right nucleus accumbens (24)<br>right lingual gyrus (21)<br>left thalamus (11)<br>left lingual gyrus (2) |
| *Amy-seeded network – anticorrelations* | | | |
| cluster 1 | +38 –26 –30 | 683 | right inferior temporal gyrus (386)<br>right temporal fusiform gyrus (142) |
| cluster 2 | –44 –30 –26 | 659 | left inferior temporal gyrus (318)<br>left temporal fusiform gyrus (133) |
| *dlPFC-seeded network – correlations* | | | |
| cluster 1 | –46 +26 +22 | 5 902 | left middle frontal gyrus (2181)<br>left frontal pole (2052)<br>left inferior frontal gyrus (663)<br>left superior frontal gyrus (169)<br>left orbitofrontal gyrus (21) |
| cluster 2 | +42 +58 +02 | 4 912 | right frontal pole (2220)<br>right middle frontal gyrus (1812)<br>right inferior frontal gyrus (158)<br>right superior frontal gyrus (91) |
| cluster 3 | +46 –50 +36 | 1 104 | right lateral occipital cortex (532)<br>right angular gyrus (477)<br>right supramarginal gyrus (13) |
| cluster 4 | –44 –60 +44 | 944 | left lateral occipital cortex (498)<br>left angular gyrus (229)<br>left supramarginal gyrus (46)<br>left superior parietal lobule (5) |
| cluster 5 | +10 +30 +44 | 170 | right dorsal anterior cingulate cortex (32)<br>right superior frontal gyrus (30)<br>left superior frontal gyrus (30)<br>left dorsal anterior cingulate cortex (16) |
| *dlPFC-seeded network – anticorrelations* | | | |
| cluster 1 | –22 –40 +76 | 2 946 | left postcentral gyrus (646)<br>right superior parietal lobule (480)<br>right postcentral gyrus (404)<br>precuneous (385)<br>left superior parietal lobule (267)<br>left precentral gyrus (131)<br>left superior frontal gyrus (30) |

| | | | |
|---|---|---|---|
| | | | right lateral occipital cortex (9) |
| cluster 2 | –06 –02 +44 | 730 | anterior cingulate gyrus (327) |
| | | | left supplementary motor area (158) |
| | | | left precentral gyrus (55) |
| | | | right supplementary motor area (39) |
| | | | right precentral gyrus (37) |
| | | | posterior cingulate cortex (14) |
| cluster 3 | –36 –16 +10 | 301 | left insular cortex (169) |
| | | | left central opercular cortex (41) |
| | | | left Heschl’s gyrus (33) |
| | | | left parietal operculum cortex (18) |
| | | | left putamen (1) |
| cluster 4 | +00 +56 +18 | 245 | left anterior cingulate cortex (60) |
| | | | right frontal pole (43) |
| | | | right anterior cingulate cortex (29) |
| | | | left frontal pole (26) |
| | | | left superior frontal gyrus (23) |
| | | | right superior frontal gyrus (16) |
| cluster 5 | +26 –06 –24 | 234 | right hippocampus (93) |
| | | | right amygdala (70) |
| | | | right parahippocampal gyrus (54) |
| *DMN – correlations* | | | |
| cluster 1 | +06 –56 +40 | 7 692 | precuneous (4235) |
| | | | posterior cingulate cortex (1561) |
| | | | lingual gyrus (286) |
| cluster 2 | +06 +60 –04 | 4 332 | frontal poles (1286) |
| | | | dorsal anterior cingulate gyri (1050) |
| | | | medial frontal cortex (512) |
| | | | anterior cingulate gyrus (422) |
| | | | superior frontal gyrus (8) |
| cluster 3 | +46 –68 +26 | 862 | right angular gyrus (629) |
| | | | right middle temporal gyrus (4) |
| cluster 4 | –42 –78 +30 | 614 | left angular gyrus (364) |
| *DMN - anticorrelations* | | | |
| cluster 1 | –46 –60 –30 | 2 251 | left cerebellum (1658) |
| | | | left occipital fusiform gyrus (115) |
| | | | left occipital pole (114) |
| | | | left temporal occipital fusiform cortex (40) |
| | | | left lateral occipital cortex (33) |
| | | | left inferior temporal gyrus (31) |
| | | | left lingual gyrus (9) |
| cluster 2 | +46 +08 +22 | 1 628 | right precentral gyrus (597) |
| | | | right inferior frontal gyrus (251) |
| | | | right temporal pole (205) |
| | | | right opercular cortex (130) |
| | | | right middle frontal gyrus (63) |

| | | | |
|---|---|---|---|
| | | | right insular (51)<br>right orbitofrontal cortex (14)<br>right planum polare (14) |
| cluster 3 | +38 –86 –04 | 1 599 | right cerebellum (903)<br>right lateral occipital cortex (397)<br>right occipital pole (163)<br>right occipital fusiform gyrus (7) |
| cluster 4 | –52 +10 –02 | 1 364 | left precentral gyrus (495)<br>left inferior frontal gyrus (257)<br>left temporal pole (184)<br>left central opercular cortex (90)<br>left frontal operculum cortex (44)<br>left middle frontal gyrus (22)<br>left insular (9)<br>left inferior frontal gyrus (9)<br>left planum polare (8)<br>left orbitofrontal cortex (4) |
| cluster 5 | +04 +04 +62 | 1 020 | supplementary motor cortex (477)<br>superior frontal gyrus (177)<br>right dorsal anterior cingulate cortex (125) |
| cluster 6 | +56 –40 +54 | 426 | right posterior supramarginal gyrus (263)<br>right anterior supramarginal gyrus (76)<br>right superior parietal lobule (19)<br>right angular gyrus (2) |
| cluster 7 | +00 –26 +10 | 112 | left thalamus (35)<br>right thalamus (13) |
| *V1-seeded network – correlations* | | | |
| cluster 1 | +02 –86 +06 | 19 801 | right lingual gyrus (1684)<br>right occipital pole (1586)<br>precuneous (1577)<br>left occipital pole (1568)<br>left lingual gyrus (1439)<br>right occipital fusiform cortex (919)<br>right calcarine cortex (894)<br>left occipital fusiform cortex (791)<br>left calcarine cortex (706)<br>right lateral occipital cortex (652)<br>right cuneal cortex (641)<br>left lateral occipital cortex (575)<br>posterior cingulate gyrus (129)<br>left parahippocampal gyrus (44)<br>right parahippocampal gyrus (8) |
| *V1-seeded network – anticorrelations* | | | |
| cluster 1 | +36 –74 –56 | 9 896 | cerebellum (4137)<br>right interior temporal gyrus (948) |

| | | |
|---|---|---|
| | | left inferior temporal gyrus (925)<br>left middle temporal gyrus (446)<br>right middle temporal gyrus (285) |

*Note.* Number of voxels per region do not add up to total cluster size due to voxels not covered by atlas labels. NAc = nucleus accumbens; Amy = amygdala; dlPFC = dorsolateral prefrontal cortex; DMN = default mode network; V1 = primary visual cortex; All clusters are significant at a FWE-corrected alpha level of $p < 0.05$.
