## Supplementary Table 2 for "Hyperconnectivity and altered dynamic interactions of a nucleus accumbens network in post-stroke depression"

Supplementary Table 2. Lesion overlap with FC networks, clusters associated with GDS scores, and white matter tracts

| | | jaccard index | | | | voxels | | | |
|---|---|---|---|---|---|---|---|---|---|
| | *n* | mean | *SD* | min | max | mean | *SD* | min | max |
| NAc-seeded | | | | | | | | | |
| correlated | 16 | 0.0063 | 0.0155 | 0 | 0.071 | 534.57 | 1339.2 | 0 | 6440 |
| anticorrelated | 3 | 0.0002 | 0.0009 | 0 | 0.005 | 6.52 | 28.13 | 0 | 170 |
| GDS cluster | 12 | 0.0104 | 0.027 | 0 | 0.107 | 236.11 | 620.834 | 0 | 2389 |
| WM tracts | 6 | 0.0002 | 0.0008 | 0 | 0.004 | 2.17 | 7.68 | 0 | 43 |
| Amy-seeded | | | | | | | | | |
| correlated | 24 | 0.0078 | 0.0132 | 0 | 0.051 | 909.5 | 1527.07 | 0 | 6078 |
| anticorrelated | 4 | 0.0021 | 0.0078 | 0 | 0.045 | 104.25 | 400.04 | 0 | 2397 |
| dlPFC-seeded | | | | | | | | | |
| correlated | 10 | 0.0011 | 0.0035 | 0 | 0.016 | 327.73 | 1009.26 | 0 | 4469 |
| anticorrelated | 9 | 0.001 | 0.004 | 0 | 0.025 | 59.41 | 254.76 | 0 | 1655 |
| DMN | | | | | | | | | |
| correlated | 10 | 0.002 | 0.0059 | 0 | 0.027 | 588.32 | 1711.6 | 0 | 8317 |
| anticorrelated | 11 | 0.0023 | 0.0072 | 0 | 0.038 | 259.05 | 845.36 | 0 | 4783 |
| GDS cluster | 4 | 0.0003 | 0.0013 | 0 | 0.007 | 31.75 | 151.33 | 0 | 868 |
| WM tracts | 4 | 0.0004 | 0.0009 | 0 | 0.005 | 1.58 | 3.9 | 0 | 21 |
| V1-seeded | | | | | | | | | |
| correlated | 8 | 0.0128 | 0.0365 | 0 | 0.174 | 2380.89 | 6876.92 | 0 | 33397 |
| anticorrelated | 6 | 0.001 | 0.0035 | 0 | 0.019 | 126.82 | 453.49 | 0 | 2592 |

*Note.* FC = functional connectivity; GDS = geriatric depression scale; SD = standard deviation; NAc = nucleus accumbens; Amy = amygdala; dlPFC = dorsolateral prefrontal-cortex; DMN = default mode network; V1 = primary auditory cortex.
